# Inflammation Beyond the Disc: Circulating Inflammatory Biomarkers in Lumbar Disc Herniation and Degeneration—A Case-Control Study

**DOI:** 10.64898/2026.08.28.26361607

**Authors:** Niroshima Dedunu Withanage, S Perera, LV Athiththan

**Author notes:** Corresponding author: E mail (NDW). These authors also contributed equally to this work (S Perera, LV Athiththan).

## Abstract

**Background:** Lumbar disc herniation, with or without concomitant disc degeneration, is a major cause of lumbar radiculopathy and low back pain, which also a key public musculoskeletal disorder without an exact pathophysiology. Studies have suggested that inflammatory cells and biochemical markers of inflammation also play an important role in lumbar radiculopathy in addition to nerve compression. The aim of the present study was to assess the association of selected circulatory inflammatory markers (CRP, hs-CRP and E-selectin) in patients with lumbar disc herniation without radiological degeneration (LDH) and lumbar disc herniation with radiological degeneration (LDHD).

**Materials & methods:** This case-control study included 208 participants, comprising 104 patients with lumbar disc pathology and 104 controls. Patients were further stratified into LDH (n=67) and LDHD (n=37). Serum CRP, hs-CRP and E-selectin concentrations were measured.

**Results:** Among the patients, 35.6 % presented with LDHD while 64.4 % had only LDH. Significantly increased median hs-CRP (p<0.001) and CRP (p<0.001) were observed in patients groups compared to controls, while CRP showing a consistent independent association across the combined disease (OR=1.68, 95% CI=1.33-2.14, p<0.001), LDHD (OR=1.62, 95% CI=1.16-2.20, p=0.005) and LDH (OR=1.69, 95% CI=1.30-2.20, p<0.001) multivariable models. No significant difference was observed in serum E-selectin between the study groups. Multivariable models incorporating inflammatory and clinical variables demonstrated substantially greater discriminatory performance than individual biomarkers alone.

**Conclusion:** Elevated circulating CRP and hs-CRP concentrations were associated with lumbar disc pathology, with CRP showing a consistent independent association across the combined disease, LDH and LDHD multivariable models, whereas E-selectin showed no significant association. Multivariable models incorporating inflammatory and clinical variables demonstrated greater discriminatory performance than individual biomarkers. These findings support a potential systemic inflammatory component in lumbar disc pathology, although the cross-sectional nature of the measurements does not establish causality or a local inflammatory response within the disc.

## Introduction

Low back pain (LBP) is considered as the major cause of disability worldwide. Among the various factors for LBP, defects associated with intervertebral disc (IVD) ranks highest such as disc herniations and disc degeneration [1–4]. However, exact pathophysiology for disc herniation or degeneration is not known. Studies have reported that (IVD) material that protrudes out the disc space, compresses the spinal nerves on either side of the spinal cord, giving rise to lumbar radiculopathy. Although this phenomenon is widely accepted, studies have shown that the pathophysiology of the exact mechanism cannot be fully explained by mechanical nerve compression alone [3,5]. Studies have suggested that inflammatory cells and biochemical markers of inflammation also play an important role in lumbar radiculopathy in addition to nerve compression [3–6]. It is further mentioned that herniation of the disc initiates the immunological and inflammatory responses and dysregulation of inflammatory state is considered as a potential mechanism in the LBP associated with disc herniation an degeneration [1]. Studies done on histology of the herniated IVDs have shown inflammatory cells on the herniated IVDs, thus leading to secrete numerous pro-inflammatory markers and cytokines including, interleukins, tumour necrosis factor (TNF)-α, intercellular adhesion molecule-1 (ICAM-1), prostaglandin E2, leukotriene B-4, thromboxane B2, phospholipase A2, nitric oxide (NO) and matrix metalloproteinases (MMPs) [2,5–8]. Among them, it is believed that IL-6 increases serum C-reactive protein (CRP) concentration [2,5,9]. When CRP level was measured by the conventional methods of immunonephlometric or immunoturbidometric assays, most patients with disc herniation had normal limits. Therefore, it was documented that such assessment of serum CRP might not detect the local inflammation around the nerve root as it occurs in a small area of disc herniation. However, with the availability of ultrasensitive latex-enhanced immunoassay for high sensitivity CRP (hs-CRP), which is useful not only in the diagnosis of myocardial infarction and stroke, but also identified as a promising marker in the detection of small inflammation around disc herniation in the lumbar spine, which can ultimately elevate systemic circulatory inflammatory markers [2].

E-selectin is a leukocyte adhesion pro-inflammatory marker which can be elevated in inflammation associated with lumbar disc herniation (LDH), and studies have proven increased levels of selectin group of adhesion molecules in many inflammatory aetiologies [3].

According to available clinical data, patients get recurrence of LBP associated with LDH even after surgical management. As patients’ responses to current treatment methods are unpredictable and patients also presented with recurrence of pain, investigators hypothesized that inflammatory pathways may contribute to lumbar disc pathology in addition to mechanical nerve compression. Although numerous studies have demonstrated inflammatory activity within herniated IVD tissue, evidence regarding circulating inflammatory biomarkers in patients with LDH remains inconsistent. Furthermore, few studies have compared patients with LDH alone and those with lumbar disc herniation and degeneration using the same panel of circulating inflammatory biomarkers. Therefore, this study investigated the association of serum CRP, hs-CRP and E-selectin concentrations with patients with lumbar disc herniation and without radiological degeneration (LDH) and patients with lumbar disc herniation and radiological degeneration (LDHD) in a Sri Lankan case-control cohort.

## Materials and methods

### Study design and setting

A case-control study was conducted with a total of 208 participants. Cases were recruited from a hospital in the capital of Sri Lanka that drains patients from all over the country and thereby represents almost all the districts of Sri Lanka. Controls represented several districts of Sri Lanka. Ethical approval was obtained from the Ethics Review Committee of the Faculty of Medical Sciences, University of Sri Jayewardenepura, Colombo, Sri Lanka (29/14). Study was conducted from May to December 2018. After detailing the study protocol, informed written consent was obtained from all participants.

### Study population

Patients who had low back pain with LDH confirmed by Magnetic Resonance Imaging (MRI) by a consultant neurosurgeon and consultant radiologist were categorized as cases (n=104). Adult volunteers without having LBP during the preceding one month of the study and, did not have LDH were categorized as control (n=104). Both case and control subjects were between 18-74 years of age. The concomitant presence of other bone disorders such as osteoarthritis, osteoporosis, pregnancy and malignancies was excluded in both cases and controls. In addition, cases with trauma and accident-related LDH were also excluded. Cases presented with herniations further stratified as, patients with lumbar disc herniation without radiological degeneration (LDH) and patients with lumbar disc herniation and with radiological degeneration (LDHD). There were 37 LDHD subjects whereas remaining cases were LDH subjects (n=67). Separate cluster analysis was conducted for combined disease groups (LHDH+LDH), LDHD and LDH.

### Sample size calculation

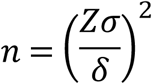

Where,

n= Sample size
Z= Standard normal deviate for chosen confidence level.
Since 95 % confidence level was used the value is 1.96
σ = Standard deviation
δ = precision of 0.5
Sample size=208

The minimum sample size was calculated using the formula for comparing two independent groups based on the expected difference in circulatory inflammatory biomarker concentrations reported in a previous study (mean hs-CRP=3mg/L; SD=3.1mg/L) [10]. Assuming a 95% confidence level (Z=1.96), a precision of 0.5, and the expected standard deviation from previous literature, the required minimum sample size was calculated as 104 participants per group. Accordingly, 104 patients and 104 controls were recruited, giving a total sample size of 208 participants.

### Specimen collection

Approximately 6 mL of venous blood sample was collected adhering to all standard precautions, under aseptic, sterile conditions from cases (before the surgery) as well as controls and serum was separated at 3000 rpm for 5 minutes and stored in −20°C for the batch analysis of circulatory inflammatory markers.

### Serum C-Reactive protein and high sensitivity C-Reactive protein analysis

Serum CRP and hs-CRP were measured using automated colourimetric method using Konelab 20 XT clinical analyser and Manufacturers reagents and kit protocols were used for the analysis. CRP standard super high calibrator, CRP control high (Biolabo, France) were used to prepare the calibration curve, while CRP HS calibrator (Thermo Scientific Co, Finland) was used for the preparation of hs-CRP calibration curve.

### Serum E-selectin analysis

Serum E-selectin was measured by ELISA technique (Sigma-Aldrich Corporation, USA) and absorbance was read at 450 nm.

### Data analysis

Statistical analyses were performed using IBM SPSS Statistics (31.0). Continuous variables were assessed for distributional characteristics. Because inflammatory biomarker concentrations were not normally distributed, data were summarized as median and interquartile range (IQR). Differences in circulatory biomarker concentrations among the control, LDH and LDHD groups were assessed using the Kruskal–Wallis test, with Dunn’s post-hoc test and Bonferroni correction for pairwise comparisons. Effect size was expressed using epsilon-squared (ε²).

Binary logistic regression was performed separately for control versus combined disease (LDH+LDHD), control versus LDH, and control versus LDHD to evaluate associations between individual circulatory biomarkers and disease status. Multivariable logistic regression models were subsequently constructed for each disease comparison after adjustment for relevant clinical and demographic variables. Results were reported as odds ratios (ORs) with 95% confidence intervals (CIs). Because CRP and hs-CRP represent closely related inflammatory measures, potential multicollinearity was assessed using Spearman correlation, tolerance and variance inflation factor (VIF). Multicollinearity was considered acceptable when VIF values were below 5 and tolerance values exceeded 0.20.

Receiver operating characteristic (ROC) curve analysis was performed to assess the discriminatory performance of individual circulatory biomarkers and the multivariable models. The area under the ROC curve (AUC) with 95% CI was reported. A two-sided p-value <0.05 was considered statistically significant.

## Results

### Characteristics of study subjects

The majority of cases were presented with LDH (64.4%) only, while others had LDHD (35.5%). Mean age of patients with LDHD, LDH and controls was 47.4±17.0, 41.5±14.8 and 43.2±15.2 years, respectively. There were more male subjects (53.7%) in cases with LDH, while there were more females (51.4%) in cases with LDHD. Both LDHD and LDH patient groups had similar BMI values (LDHD=27.4±4.9 kg/m^2^, LDH=27.5±5.6 kg/m^2^).

### Association of inflammatory markers with LDH and LDHD subjects

Comparison of inflammatory biomarkers among the three study groups demonstrated a significant difference in serum hs-CRP concentrations (Kruskal–Wallis H = 15.62, p < 0.001), with a small effect size (ε²=0.06). Serum CRP concentrations also differed significantly among the groups, with a similarly small effect size (ε²=0.07). In contrast, serum E-selectin concentrations did not differ significantly among the study groups (p > 0.05). Although the highest median E-selectin concentration was observed in the control group [median=13.35 (2.62–21.82) ng/mL], the effect size was negligible (ε² < 0.001) (Table 1).

**Table 1:** Association of selected inflammatory markers between study groups.

|  | **Control**<br>**group**<br>**Median**<br>**(Q1-Q3)**<br>**n=104** | **LDHD**<br>**subgroup**<br>**Median**<br>**(Q 1-Q3)**<br>**n=37** | **LDH**<br>**subgroup**<br>**Median**<br>**(Q1-Q3)**<br>**n=67** | **H**<br>**value** | **Kruskal-**<br>**Wallis**<br>**p value** | **$\epsilon^2$** |
| --- | --- | --- | --- | --- | --- | --- |
| hs-CRP (mg/L) | 0.88<br>(0.23-1.62) | 1.63<br>(0.59-2.81) | 1.59<br>(0.70-4.46) | 15.62 | <0.001* | 0.06 |
| CRP (mg/L) | 3.00<br>(1.82-4.27) | 4.00<br>(0.25-5.85) | 4.30<br>(2.90-7.30) | 14.43 | <0.001* | 0.07 |
| E-selectin<br>(ng/mL) | 13.35<br>(2.62-21.82) | 7.90<br>(3.80-19.50) | 7.30<br>(5.10-17.10) | 0.046 | 0.977 | <0.001 |

Kruskal-Wallis analysis, values presented as median (IQR), \*p<0.001 considered as significant, $\epsilon^2$ -effect size,

Post hoc pairwise comparisons using Dunn’s test with Bonferroni correction demonstrated that serum hs-CRP concentrations were significantly higher in patients with LDH than in healthy controls (adjusted p<0.001). However, no significant differences were observed between the control and LDHD groups (adjusted p=0.068) or between the LDH and LDHD groups (adjusted p=1.000). Similarly, serum CRP concentrations were significantly higher in both the LDHD (adjusted p=0.040) and LDH (adjusted p=0.001) groups than in controls, while no significant difference was observed between the two patient groups (adjusted p=1.000).

In binary logistic regression, higher hs-CRP (OR=1.09, 95% CI=1.02-1.19, p=0.018)) and CRP (OR=1.17, 95% CI=1.06-1.29, p=0.002) concentrations were positively associated with the combined disease group compared with controls. CRP was also significantly associated with both LDHD (OR=1.12, 95% CI=1.01-1.25, p=0.041) and LDH (OR=1.18, 95% CI=1.07-1.30, p=0.001) when analyzed separately, whereas hs-CRP showed a significant association with LDH (OR=1.11, 95% CI=1.03-1.21, p=0.011) but not with LDHD. E-selectin was not significantly associated with disease status in any of the three models (Table 2: a, b, c).

**Table 2:**
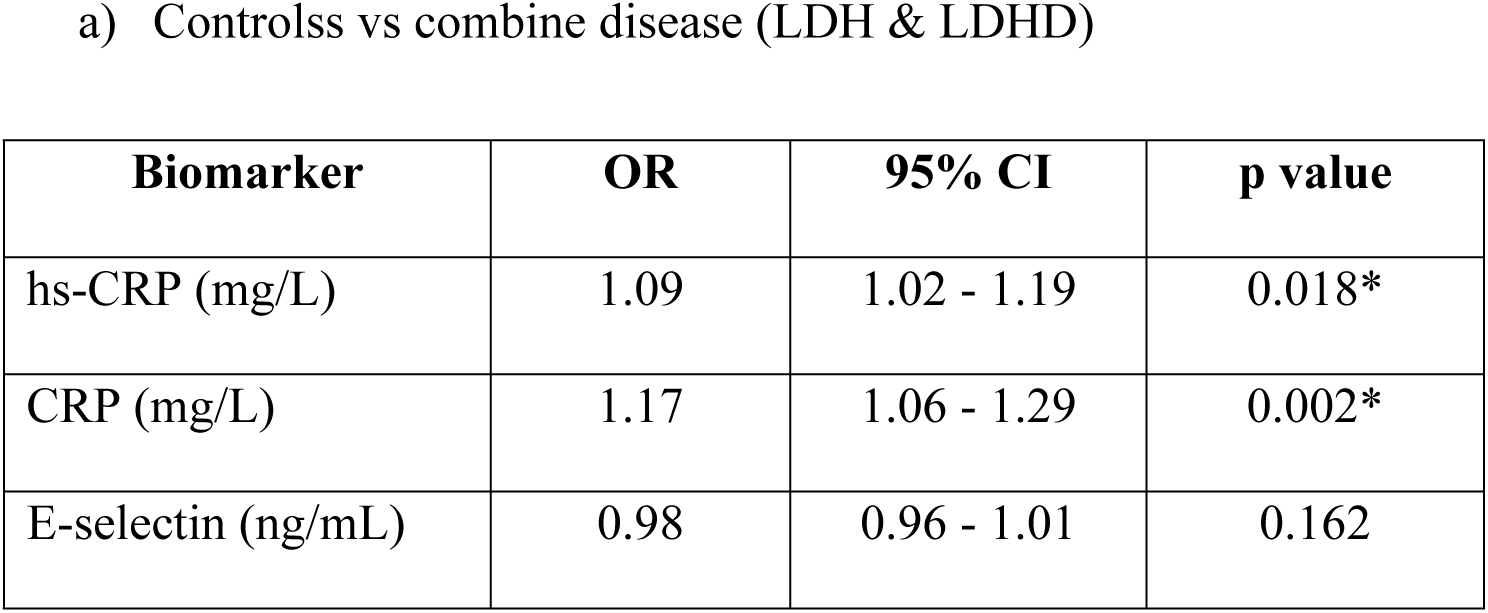
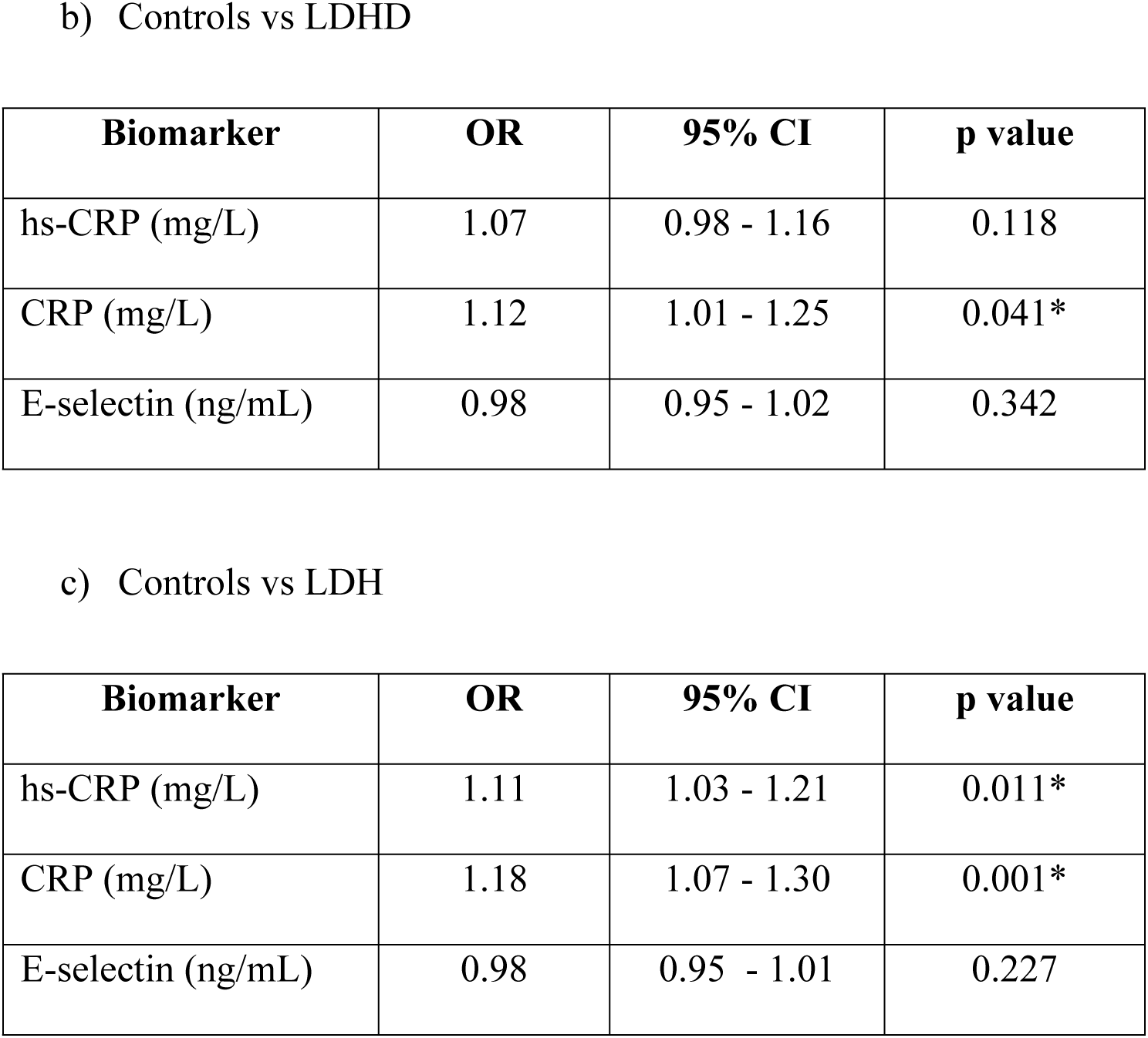
Association of individual inflammatory biomarkers with disease status.

a) Controlss vs combine disease (LDH & LDHD)
| **Biomarker** | **OR** | **95% CI** | **p value** |
| --- | --- | --- | --- |
| hs-CRP (mg/L) | 1.09 | 1.02 - 1.19 | 0.018* |
| CRP (mg/L) | 1.17 | 1.06 - 1.29 | 0.002* |
| E-selectin (ng/mL) | 0.98 | 0.96 - 1.01 | 0.162 |

b) Controls vs LDHD
| **Biomarker** | **OR** | **95% CI** | **p value** |
| --- | --- | --- | --- |
| hs-CRP (mg/L) | 1.07 | 0.98 - 1.16 | 0.118 |
| CRP (mg/L) | 1.12 | 1.01 - 1.25 | 0.041* |
| E-selectin (ng/mL) | 0.98 | 0.95 - 1.02 | 0.342 |

c) Controls vs LDH
| **Biomarker** | **OR** | **95% CI** | **p value** |
| --- | --- | --- | --- |
| hs-CRP (mg/L) | 1.11 | 1.03 - 1.21 | 0.011* |
| CRP (mg/L) | 1.18 | 1.07 - 1.30 | 0.001* |
| E-selectin (ng/mL) | 0.98 | 0.95 - 1.01 | 0.227 |

Multivariable models demonstrated substantially greater discriminatory performance than individual inflammatory biomarkers. The model differentiating controls from the combined disease group demonstrated an AUC of 0.882 (95% CI: 0.834–0.930; p<0.001). Similarly, the models differentiating controls from LDHD and LDH demonstrated AUCs of 0.872 (95% CI: 0.822–0.922; p<0.001) and 0.874 (95% CI: 0.824–0.924; p<0.001), respectively (Table 4). CRP and hs-CRP demonstrated moderate correlation (Spearman’s ρ=0.529, p<0.001). Collinearity diagnostics showed VIF values of 4.18 for CRP and 4.22 for hs-CRP, indicating moderate but not severe multicollinearity. Both biomarkers were therefore retained in the multivariable models because they represent clinically relevant inflammatory measures with potentially distinct analytical characteristics.

**Table 3:** Multivariable analysis of inflammatory biomarkers and disease status.

| Comparison | Biomarker | Adjusted<br>OR | 95% CI | p |
| --- | --- | --- | --- | --- |
| Controls vs<br>combine disease | Age | 1.02 | 0.99 - 1.05 | 0.204 |
|  | BMI | 1.28 | 1.13 - 1.44 | <0.001* |
|  | Gender | 2.05 | 0.87 - 4.83 | 0.101 |
|  | hs-CRP | 0.79 | 0.64 - 0.93 | 0.006* |
|  | **CRP** | **1.68** | **1.33 - 2.14** | **<0.001\*** |
|  | Vitamin D | 0.71 | 0.63 - 0.82 | <0.001* |
| Controls vs LDHD | Age | 1.05 | 1.01 - 1.10 | 0.02 |
|  | BMI | 1.25 | 1.08 - 1.45 | 0.002* |
|  | Gender | 2.33 | 0.70 - 7.75 | 0.167 |
|  | hs-CRP | 0.74 | 0.54 - 1.01 | 0.056 |
|  | **CRP** | **1.62** | **1.16 - 2.26** | **0.005\*** |
|  | Vitamin D | 0.69 | 0.58 - 0.83 | <0.001* |
| Controls vs LDH | Age | 0.99 | 0.96 - 1.03 | 0.630 |
|  | BMI | 1.32 | 1.14 - 1.53 | <0.001* |
|  | Gender | 1.85 | 0.66 - 5.16 | 0.239 |
|  | hs-CRP | 0.79 | 0.65 - 0.97 | 0.027 |
|  | **CRP** | **1.69** | **1.30 - 2.20** | **<0.001\*** |
|  | Vitamin D | 0.69 | 0.59 - 0.82 | <0.001* |

**Table 4:** ROC analysis.

| Comparison | Biomarker | AUC | 95% CI | p |
| --- | --- | --- | --- | --- |
| Controls vs<br>combine dsisease | hs-CRP | 0.656 | 0.582–0.729 | <0.001* |
|  | CRP | 0.652 | 0.577–0.726 | <0.001* |
|  | E-selectin | 0.501 | 0.420–0.583 | 0.975 |
|  | Multivariable model | 0.882 | 0.834–0.930 | <0.001* |
| Controls vs LDHD | hs-CRP | 0.630 | 0.527–0.732 | 0.013* |
|  | CRP | 0.639 | 0.532–0.746 | 0.011* |
|  | E-selectin | 0.498 | 0.398–0.599 | 0.976 |
|  | Multivariable model | 0.872 | 0.822–0.922 | <0.001* |
| Controls vs LDH | hs-CRP | 0.670 | 0.589–0.751 | <0.001* |
|  | CRP | 0.659 | 0.573–0.745 | <0.001* |
|  | E-selectin | 0.503 | 0.416–0.590 | 0.949 |
|  | Multivariable model | 0.874 | 0.824–0.924 | <0.001* |

AUC = area under the receiver operating characteristic curve, Multivariable model includes age, BMI, sex, vitamin D, CRP and hs-CRP

## Discussion

LDH and LDHD is considered as an important public health problem worldwide because of its enormous impact on socio-economic burden of the country. It has also been reported by Urban and Roberts (2003) that the prevalence of LDHD could be between 12-35 % [11].

Acute phase proteins such as hs-CRP and CRP are elevated in response to inflammation. These markers are regarded as most sensitive markers of inflammation [12–14]. It has been reported that LDH and LDHD induce a circulatory inflammatory response around the nerve roots which cause the pain associated with LDH and LDHD [8, 15–19]. Furthermore, evidence also suggest herniated and degenerated disc cells produce pro-inflammatory and inflammatory markers such as cytokines, interleukins (IL-1, IL-6 and IL-8), prostaglandin E_2_, TNF-α, thromboxane B_2_, MMPs and NO [8, 15]. E-selectin is also regarded as a circulatory inflammatory marker and its role as a systemic inflammatory marker in rheumatoid arthritis and LDH has been documented [3, 20, 21].

Previous studies have suggested that inflammatory pathways may contribute to symptoms associated with LDH and LDHD in addition to mechanical nerve compression [2,3,8,16]. Hence, the present study was conducted to investigate the association of selected circulatory inflammatory markers such as hs-CRP, CRP and E-selectin with LDH and LDHD with comparison to controls.

According to the analysis, the median concentrations of hs-CRP and CRP in patients with LDH and LDHD were higher than the concentrations observed in controls, showing a significant difference between the study groups for hs-CRP and CRP (p<0.001 and p<0.001).

The levels of hs-CRP and CRP reported in our study are in accordance with several previous studies. Sugimori et al (2003) reported a similar observation. Findings of the study revealed that mean hs-CRP concentration in patients with LDH (0.056 ± 0.076 mg/dL) was significantly higher (p=0.006) than in control subjects (0.017±0.021 mg/dL) and the authors recorded normal hs-CRP levels in both cases and controls. However, they were unable to correlate the increased concentration of hs-CRP with the level and type of herniation. Authors of the above study have concluded that hs-CRP might be elevated as an inflammatory response to the compression of the nerve root followed by disc herniation [15].

Another supported study finding observed that ultra-sensitive CRP levels in patients with sciatica (1.64 mg/L) were found to be significantly higher (p=0.002) than the control group (0.74 mg/L) [22]. Interestingly, a strong association between pain and hs-CRP levels in patients with acute sciatic pain was reported by Strümmer et al (2005). However, they were unable to find a relationship between hs-CRP and chronic low back pain [2]. A notable source of support for our findings was reported by Ackerman and Zhang (2006) where elevation in hs-CRP levels by percentages of 0 %, 20 %, 80 % and 73 % were observed in patients suffering from lumbar disc protrusion, prolapse, extrusion and sequestered types of herniations respectively [23].

In contrast to our findings, a study has highlighted normal hs-CRP values (1.1 mg/L) in patients with LBP or radiculopathy due to LDH, spinal stenosis and facet syndrome [9]. Similarly, another group of investigators also argued that hs-CRP could not provide enough evidence to support the relationship of hs-CRP with acute and chronic LBP [24].

Evidence suggests that prolapsed or degenerated discs can induce the expression of macrophages, which release cytokines, especially IL-6, that trigger the secretion of CRP [2,13,16]. As hs-CRP and CRP can be elevated in other inflammatory and cardiac diseases, we have excluded patients with acute or chronic infection/inflammation and with any abnormalities in electrocardiogram, which further validate the results obtained in the present study.

Among the evaluated inflammatory biomarkers, CRP demonstrated the most consistent independent association across all multivariable disease-status models in the present study which remains the major emphasis in the study. Therefore, our findings provide evidence of an association between circulating hs-CRP/CRP concentrations and lumbar disc pathology, although the observational design does not establish causality or determine whether these circulating markers reflect local inflammatory processes within the disc.

The present study did not show a significant difference in E-selectin levels between the study groups. A notable supportive finding similar to our study was reported by a study conducted in older patients (60 years and older) with LBP. This study reported no association between E-selectin and pain or pain-related functions in LBP. The study further mentioned that the above finding on E-selectin in LBP might be due to the chronic state of disc degeneration in older patients and also due to the exclusion of patients with radicular pain [20].

In contrast to our finding, another reported study with LDH patients has highlighted that cases with positive straight raise test have higher rates of immunostaining with E-selectin present on the herniated disc. The authors further suggested that E-selectin antagonist therapy can be beneficial in patients with LDH and LDHD to reduce pain, as authors argue the pain associated could be due to high E-selectin level on the IVD [21]. A similar theme of observation was noted in patients with herniated lumbar discs (n=50) [3]. Most of the reported work on E-selectin studies has focused on the immunostaining of E-selectin in the herniated IVDs, the discrepancy of the present study may partly reflect the different biological compartments assessed, as previous studies have predominantly evaluated E-selectin expression within disc tissue whereas the present study measured circulating serum E-selectin.

Interestingly, CRP demonstrated a consistent independent association across the combined disease, LDH-specific and LDHD-specific multivariable models after adjustment for age, sex, BMI and vitamin D. In contrast, the adjusted association of hs-CRP was attenuated and changed direction after adjustment. Given the moderate correlation observed between CRP and hs-CRP (Spearman’s ρ= 0.529, VIF ≈4.2), the adjusted hs-CRP coefficient likely represents the residual association after accounting for information shared with CRP rather than a biologically protective effect. Therefore, the adjusted hs-CRP estimates should be interpreted cautiously. Further, the ROC models demonstrated good apparent discriminatory performance within the study cohort, though an external validation is required before clinical application.

However, investigators identified several limitations of the present study. First, the case-control design limits causal inference and does not establish whether increased circulating inflammatory markers precede or result from lumbar disc pathology. Second, biomarkers were measured in peripheral serum and therefore may not directly reflect the inflammatory microenvironment within the intervertebral disc. Third, although participants with clinically apparent inflammatory and malignant conditions were excluded, residual confounding by unmeasured inflammatory, metabolic, lifestyle and clinical factors cannot be excluded. Fourth, the study was conducted at a single clinical centre, which may limit generalizability. Finally, the multivariable models and their discriminatory performance require validation in independent cohorts before their clinical utility can be established.

## Conclusion

This case-control study demonstrated significant associations between circulating CRP and hs-CRP concentrations and lumbar disc pathology, with CRP showing a consistent independent association across the combined disease, LDH and LDHD multivariable models. In contrast, circulating E-selectin was not significantly associated with disease status. Multivariable models incorporating inflammatory and clinical variables demonstrated substantially greater discriminatory performance than individual biomarkers alone suggesting that circulating inflammatory markers may contribute to distinguishing patients with lumbar disc pathology from controls. These findings support a potential systemic inflammatory component in lumbar disc pathology. However, because inflammatory markers were measured in peripheral serum and the study employed a case-control design, the findings do not establish causality or confirm local inflammatory activity within the intervertebral disc. Further, longitudinal studies integrating circulating biomarkers with tissue-based inflammatory profiling are warranted to clarify the role of systemic inflammation in lumbar disc pathology.

## Data Availability

All relevant data from this study will be made publicly availble upon study completion and if needed upon request by the editors, data can be produced.

## Supporting information

S1 File. Multivariable regression data

